# Geographic use and performance of distributed home video-electroencephalography in Australia

**DOI:** 10.64898/2026.09.07.26362474

**Authors:** Ewan S. Nurse, Dean R. Freestone, Emma Foster, Gabriel Dabscheck, Patrick Kwan, Mark J. Cook

## Abstract

**Objective:** To determine the geographic use of a distributed Australian home video-electroencephalography (video-EEG) service and whether diagnostic yield or technical performance varied with remoteness, clinic distance, or area-level socioeconomic disadvantage.

**Methods:** We retrospectively analysed 3,502 home video-EEG studies from 3,457 patients recorded at 24 clinic locations between September 2022 and March 2024. Patient postcodes were linked to the Modified Monash Model (MMM) and the Index of Relative Socio-economic Disadvantage (IRSD). Straight-line distance to the assigned clinic was compared with distance to the nearest public comprehensive epilepsy centre. Modified Poisson and linear models examined event, diagnostic, video, and final-check impedance outcomes.

**Results:** Studies originated from 1,112 postcodes; 854 (24.4%) were regional/rural and 17 (0.5%) remote/very remote. For regional/rural studies, the assigned clinic was a median 59.3 km closer (interquartile range [IQR]: 1.2 km farther to 116.6 km closer) than the public-centre comparator and was closer in 68.0% of studies. Median referral-to-recording time was 33.9 days in regional/rural studies and 25.5 days in metropolitan studies. At least one reported or discovered event occurred in 54.7% of studies. Adjusted analyses showed no evidence that remoteness was associated with poorer event ascertainment, seizure or interictal findings, video observability, or final-check impedance. No clinic-distance or socioeconomic association remained significant after multiplicity correction.

**Significance:** Distributed home video-EEG brought the point of connection closer to many regional/rural recipients without evidence of lower diagnostic yield or poorer technical performance among service recipients. Regional/rural referral-to-recording intervals were longer, and remote/very remote representation was sparse, so comparable access gains in genuinely remote populations remain uncertain.

**Key Points:**

- A national home video-electroencephalography service was used across 1,112 postcodes; 24.4% of studies were regional/rural.
- For regional/rural studies, assigned clinics were 59.3 km closer (interquartile range: 1.2 km farther to 116.6 km closer).
- Remoteness and clinic distance were not associated with lower diagnostic yield or poorer technical performance.
- Regional/rural referral-to-recording time was 8.4 days longer than in metropolitan studies.
- Remote/very remote data were sparse, so comparable access gains in genuinely remote populations remain uncertain.

## Introduction

Prolonged video-electroencephalography (video-EEG) is an established diagnostic tool when routine assessment is insufficient. Conventional inpatient monitoring is resource intensive and depends on specialist infrastructure.^1,2^ Minimum technical requirements for ambulatory electroencephalography (EEG) have been defined,^3^ and large clinical cohorts have shown substantial diagnostic yield with high event and video capture from home video-EEG.^4–6^ A remaining service question is whether this model can be delivered at geographic scale without compromising diagnostic yield or technical performance.

Access is particularly relevant in Australia, where specialist epilepsy services are concentrated in major cities and regional patients may face substantial travel to access EEG.^7–9^ Seizure-related hospitalisation hotspots have also been identified more often in regional and remote areas, including locations with limited access to specialised services.^10^ These patterns make the location of prolonged EEG services an important component of access, but do not by themselves show the geographic distribution of use.

Geographic decentralisation could also affect recording performance if patients farther from a clinic have less access to electrode maintenance or technical support. Prior work found no association between residential remoteness and electrode impedance during prolonged home recording,^11^ and a national cohort showed high EEG and video availability during captured events.^4^ It remains unclear whether locality, clinic distance, or area-level socioeconomic disadvantage are associated with diagnostic yield or technical performance across a distributed service.

We therefore examined an Australia-wide cohort of patients who received home video-EEG. We first characterised the geographic distribution of service use and its travel advantage relative to the nearest public comprehensive epilepsy centre. We then tested whether locality, clinic distance, or area-level disadvantage were associated with event ascertainment, diagnostic findings, or recording quality.

## Materials and Methods

### Study design and setting

This retrospective cohort study used clinical data from consecutive patients referred to a national Australian ambulatory video-electroencephalography (video-EEG) service between September 2022 and March 2024 (Seer Medical). Patients were seen at 24 EEG clinic locations (14 metropolitan, 10 regional/rural, 0 remote/very remote) for diagnostic evaluation, event or seizure classification, treatment assessment, or presurgical evaluation. Regional/rural clinic locations operated periodically rather than as continuously staffed permanent sites. We linked clinical and referral data to neurologist reports, remotely logged electrode impedance, and postcode-derived geography. Primary video-EEG recordings were not re-reviewed.

Studies were eligible when a neurologist report was available and recording duration was 3 hours to 14 days. Repeated studies were retained and linked by patient identifier. The St Vincent’s Hospital Melbourne Human Research Ethics Committee approved the study (project 57392), and written informed consent was provided by patients or, where applicable, a guardian. Reporting followed the Strengthening the Reporting of Observational Studies in Epidemiology statement.^12^

### Ambulatory video-EEG monitoring

The clinical service and recording system have been described previously.^4,11^ EEG was recorded at 250 samples per second using a full 10-20 montage with 21 recording electrodes and simultaneous electrocardiography (ECG); all studies included concurrent video.^3^ Recordings were initiated in a clinic and completed at home. Patients or carers logged events using a mobile application or paper diary. During acquisition, trained technicians remotely reviewed EEG and video quality, electrode impedance, and system status and provided technical support when required. A neurophysiology scientist and neurologist subsequently reviewed the recording, and the neurologist issued the final clinical report.

### Locality classification

Patient postcode was linked to the Australian Government Modified Monash Model (MMM) 2019 classification.^13^ MMM 1 was classified as metropolitan, MMM 2-5 as regional/rural, and MMM 6-7 as remote/very remote; where a postcode contained multiple MMM values, the least remote was assigned. Postcode centroids were also linked to the 2021 Australian Bureau of Statistics Index of Relative Socio-economic Disadvantage (IRSD), with higher scores indicating less disadvantage.^14^ Straight-line distance was calculated from the residential postcode centroid to the assigned clinic and, as a contextual comparator, to the nearest public comprehensive epilepsy centre in the Australian Institute of Health and Welfare MyHospitals inventory.^15^ These measures use postcode centroids rather than observed road travel; the public-centre comparator does not represent an observed alternative referral destination. For descriptive context, the geographic distribution of the Australian population was obtained from Australian Institute of Health and Welfare national estimates using the separate Remoteness Area classification; these categories are not directly equivalent to MMM groups.

### Clinical variables and outcomes

Clinical covariates extracted from the database included age, self-reported sex, epilepsy and disability status, study duration, referral details, and current medications. Referral-to-recording interval was calculated from the linked referral date to recording start. Referral purpose was classified as initial diagnosis, event or seizure classification, treatment response or seizure burden, presurgical evaluation, or other/unclear.

Events logged by patients or carers were classified as reported, and additional events identified during clinical review were classified as discovered. Study-level event outcomes were any reported or discovered event, any reported event, and any discovered event. Diagnostic findings were derived from the neurologist report.

Recording-quality outcomes were derived from the neurologist report. Major acquisition or interpretability issues comprised EEG interpretation limited by artefact, electrode disconnection/removal/channel loss, EEG interruption/data loss, or an event without usable video. Video unavailability could be technical (e.g., camera or recording failure), contextual (e.g., an event occurred when video recording was not practically available), or intentional (e.g., video recording had been disabled). Artefact or noise without a major issue was classified separately, as were events partly or fully outside the camera view. These endpoints indicate that an issue occurred at some point during a study, not that the entire recording was unusable.

For the impedance analysis, we used the last impedance measurement on the final study day and calculated the percentage of the 21 recording electrodes below 10 kΩ.^3^

### Statistical analysis

Continuous variables are reported as median (interquartile range [IQR]) and categorical variables as n/N (%). Unadjusted comparisons used Kruskal-Wallis tests for continuous outcomes and Pearson chi-square, Fisher exact, or 100,000-sample fixed-margin Monte Carlo exact tests for categorical outcomes as appropriate. Benjamini-Hochberg correction controlled the false-discovery rate within prespecified test families.

MMM associations with binary event, diagnostic, and video outcomes were estimated by modified Poisson regression with robust variance, adjusted for age, sex, recorded epilepsy diagnosis, disability, antiseizure medication count, referral purpose, and recording duration; covariance was clustered by patient and postcode. Metropolitan studies were the reference. Sparse endpoints that did not support stable two-way-clustered models were assessed by exact tests.

Final-check impedance used linear regression adjusted for age, sex, recorded epilepsy diagnosis, disability, and recording duration with the same clustering. Geographic-gradient models entered log2-transformed assigned-clinic distance and standardised IRSD together in the corresponding binary or impedance model. Effects were interpreted per doubling of distance and per 1-standard deviation (SD) higher IRSD; false-discovery-rate correction was applied across the 10 binary tests and separately across the two impedance tests.

Relative distance was summarised as assigned-clinic minus public-centre distance and as the assigned/public-centre distance ratio. An exploratory clustered linear model of log2 ratio included MMM group and IRSD. In 5,000 sensitivity simulations, postcode-centroid origins were displaced within maximum radii of 2.5 km (metropolitan), 25 km (regional/rural), and 75 km (remote/very remote) before distances were recalculated.

To assess sensitivity to clinic-level differences, the adjusted MMM and geographic-gradient outcome models were repeated with assigned clinic included as a categorical fixed effect while retaining patient- and postcode-clustered covariance. Clinic-locality overlap was summarised to assess the information available for within-clinic MMM comparisons. Referral-to-recording interval was examined separately in an exploratory clinic-adjusted model.

Bayes factors were complementary. BF01, favouring models without MMM, was approximated from Bayesian information criterion differences between unclustered models with and without MMM terms.^16^ Comparisons used covariate-matched logistic models for binary endpoints, covariate-matched linear models for final-check impedance, and unadjusted linear models for referral-to-recording interval. Bayes factors were not primary inferential tests. All tests were two-sided; analyses used Python 3.11 and statsmodels.^17^

## Results

### Cohort and locality distribution

The primary analysis included 3,502 ambulatory video-EEG studies from 3,457 patients recorded from September 2022 to March 2024. The cohort comprised 2,631 metropolitan studies (75.1%), 854 regional/rural studies (24.4%), and 17 remote/very remote studies (0.5%) across 1,112 unique patient postcodes (Table 1; Figure 1). Median age was 33 years (IQR 21-53), and median concurrent antiseizure medication count was 0 (IQR 0-1, range 0-7). Initial diagnosis was the most common referral purpose (44.8%), followed by treatment response or seizure burden (28.8%) and event or seizure classification (17.8%); presurgical and other/unclear indications accounted for 8.6%.

**Figure 1.**
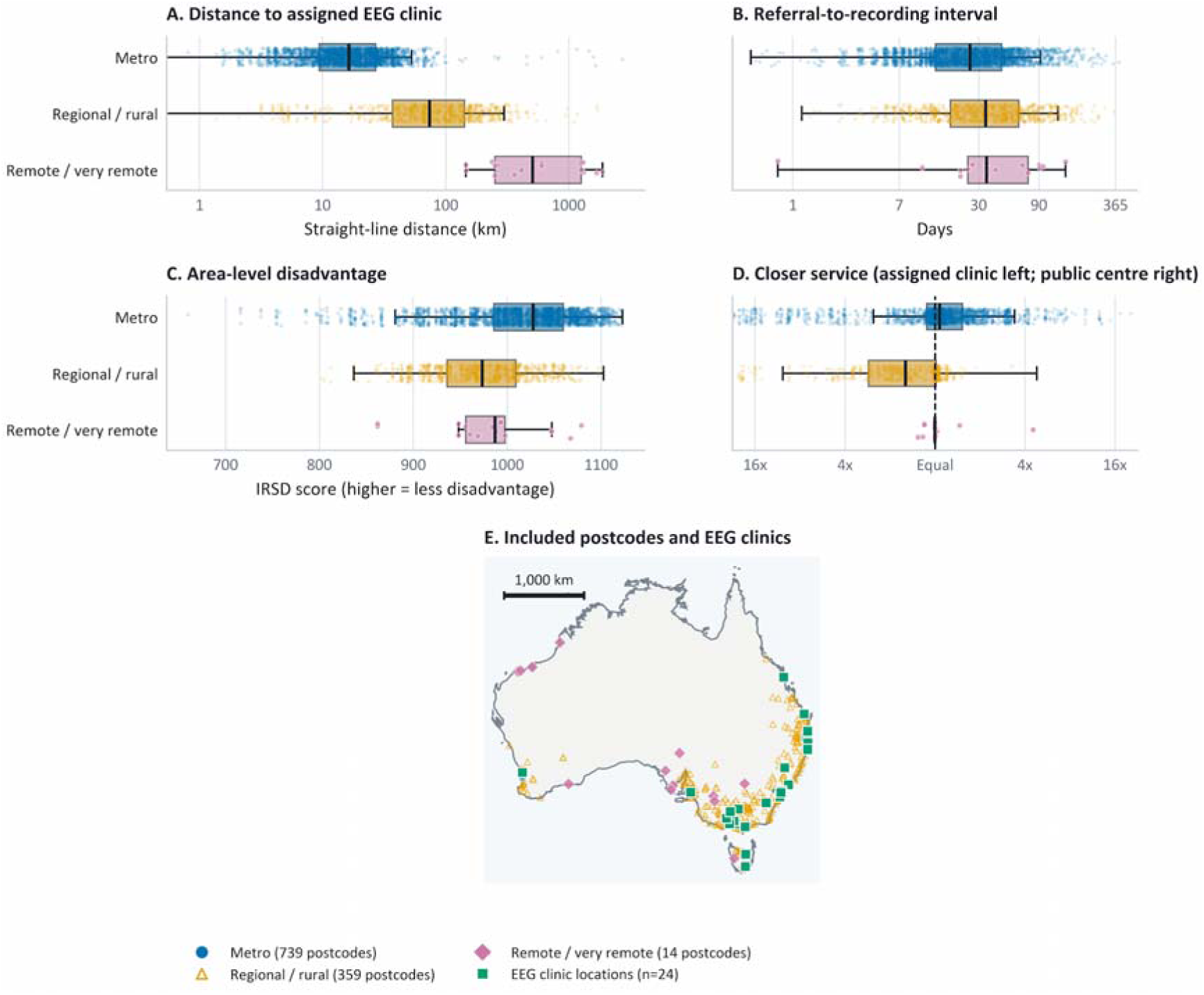
Geographic use, referral-to-recording interval, and clinic distribution of home video-EEG. (A) Distance to the assigned EEG clinic by MMM group. (B) Referral-to-recording interval by MMM group, calculated from linked referral date to recording start. (C) Area-level IRSD by MMM group. For panels A-C, points are individual studies, boxes show medians and IQRs, and whiskers extend to 1.5 times the IQR. Referral-to-recording interval differed by MMM group (Kruskal-Wallis P < 0.001); medians were 25.5 days in metropolitan and 33.9 days in regional/rural studies. (D) Assigned-clinic distance compared with distance to the nearest AIHW-listed public comprehensive epilepsy centre. Points left of Equal favour the assigned clinic; points right of Equal favour the public centre, and tick labels show the fold difference. (E) Distribution of unique included patient postcodes and the 24 EEG clinic locations. Distances are straight-line postcode-centroid measures, not observed road travel. AIHW centres are geographic comparators only; the inventory excludes private providers and is not age-specific. AIHW, Australian Institute of Health and Welfare; EEG, electroencephalography; IQR, interquartile range; IRSD, Index of Relative Socio-economic Disadvantage; MMM, Modified Monash Model.

**Table 1.** Cohort characteristics by MMM group. EEG, electroencephalography; IQR, interquartile range; IRSD, Index of Relative Socio-economic Disadvantage; MMM, Modified Monash Model.

| Characteristic | Overall | Metro | Regional / rural | Remote / very remote |
| --- | --- | --- | --- | --- |
| Studies, n | 3,502 | 2,631 | 854 | 17 |
| Patients, n | 3,457 | 2,592 | 848 | 17 |
| Age, years, median [IQR] | 33.0 [21.0, 53.0] | 32.0 [21.0, 53.0] | 34.0 [20.0, 53.8] | 21.0 [12.0, 45.0] |
| Concurrent antiseizure medications, median [IQR] | 0.0 [0.0, 1.0] | 0.0 [0.0, 1.0] | 0.0 [0.0, 1.0] | 0.0 [0.0, 1.0] |
| Distance to assigned EEG clinic, km, median [IQR] | 19.8 [10.3, 38.3] | 16.4 [9.4, 27.0] | 73.9 [36.9, 142.0] | 509.6 [250.2, 1261.3] |
| Area-level IRSD score, median [IQR] | 1011.5 [965.8, 1050.0] | 1027.7 [985.7, 1059.7] | 973.7 [936.0, 1009.1] | 987.2 [955.7, 997.7] |
| Sex |  |  |  |  |
| Male | 1,422/3,485 (40.8%) | 1,069/2,616 (40.9%) | 348/852 (40.8%) | 5/17 (29.4%) |
| Female | 2,063/3,485 (59.2%) | 1,547/2,616 (59.1%) | 504/852 (59.2%) | 12/17 (70.6%) |
| Unknown/missing | 17/3,502 (0.5%) | 15/2,631 (0.6%) | 2/854 (0.2%) | 0/17 (0.0%) |
| Epilepsy diagnosis |  |  |  |  |
| Yes | 1,653/2,921 (56.6%) | 1,248/2,213 (56.4%) | 398/697 (57.1%) | 7/11 (63.6%) |
| No | 1,268/2,921 (43.4%) | 965/2,213 (43.6%) | 299/697 (42.9%) | 4/11 (36.4%) |
| Unknown/missing | 581/3,502 (16.6%) | 418/2,631 (15.9%) | 157/854 (18.4%) | 6/17 (35.3%) |
| Disability |  |  |  |  |
| Yes | 830/3,498 (23.7%) | 596/2,628 (22.7%) | 233/853 (27.3%) | 1/17 (5.9%) |
| No | 2,668/3,498 (76.3%) | 2,032/2,628 (77.3%) | 620/853 (72.7%) | 16/17 (94.1%) |
| Unknown/missing | 4/3,502 (0.1%) | 3/2,631 (0.1%) | 1/854 (0.1%) | 0/17 (0.0%) |

Median straight-line distance to the assigned EEG clinic was 16.4 km (IQR 9.4-27.0) in metropolitan studies, 73.9 km (IQR 36.9-142.0) in regional/rural studies, and 509.6 km (IQR 250.2-1261.3) in remote/very remote studies. Among regional/rural studies with paired geographic measures, median distance to the nearest Australian Institute of Health and Welfare (AIHW)-listed public comprehensive epilepsy centre was 148.8 km (IQR 91.5-235.3). The paired assigned-clinic minus public-centre distance was −59.3 km (IQR −116.6 to 1.2), favouring the assigned clinic, which was closer in 68.0% of studies. This median one-way difference corresponds to 118.6 km for a return journey and 237.2 km across the four one-way journey legs typically required for connection and disconnection. Among remote/very remote studies, the paired difference was −2.2 km (IQR −6.6 to 1.7), with the assigned clinic closer in 11/17 studies (64.7%) (Table S1).

Referral-to-recording intervals differed by MMM group (Kruskal-Wallis P < 0.001; false-discovery-rate q = 0.001). Median intervals were 25.5 days (IQR 13.5-45.5) in metropolitan studies, 33.9 days (IQR 17.8-62.3) in regional/rural studies, and 34.6 days (IQR 24.5-73.7) in remote/very remote studies. The regional/rural-metropolitan difference in medians was 8.4 days. In an exploratory model additionally adjusting for assigned clinic, the regional/rural difference was no longer evident (ratio estimate 0.99; 95% CI 0.91-1.07; P = 0.81; Table S5). Median area-level IRSD scores were 1027.7, 973.7, and 987.2, respectively.

### Event ascertainment and diagnostic findings

At least one reported or discovered event was present in 1,915 of 3,502 studies (54.7%). The proportion was 54.3% in metropolitan studies, 55.9% in regional/rural studies, and 58.8% in remote/very remote studies. At least one patient- or carer-reported event was present in 50.1% of studies, and at least one event discovered during clinical review was present in 12.3%. Epileptic seizures were identified in 455 studies (13.0%), and an interictal abnormality was documented in 2,156 studies (61.6%) (Figure 2). Adjusted modified Poisson models showed no evidence of an association between MMM group and any event or diagnostic endpoint (Table S2). For any event, adjusted prevalence ratios (PRs) were 1.01 (95% confidence interval [CI] 0.94-1.08) for regional/rural studies and 1.09 (95% CI 0.77-1.54) for remote/very remote studies relative to metropolitan studies.

**Figure 2.**
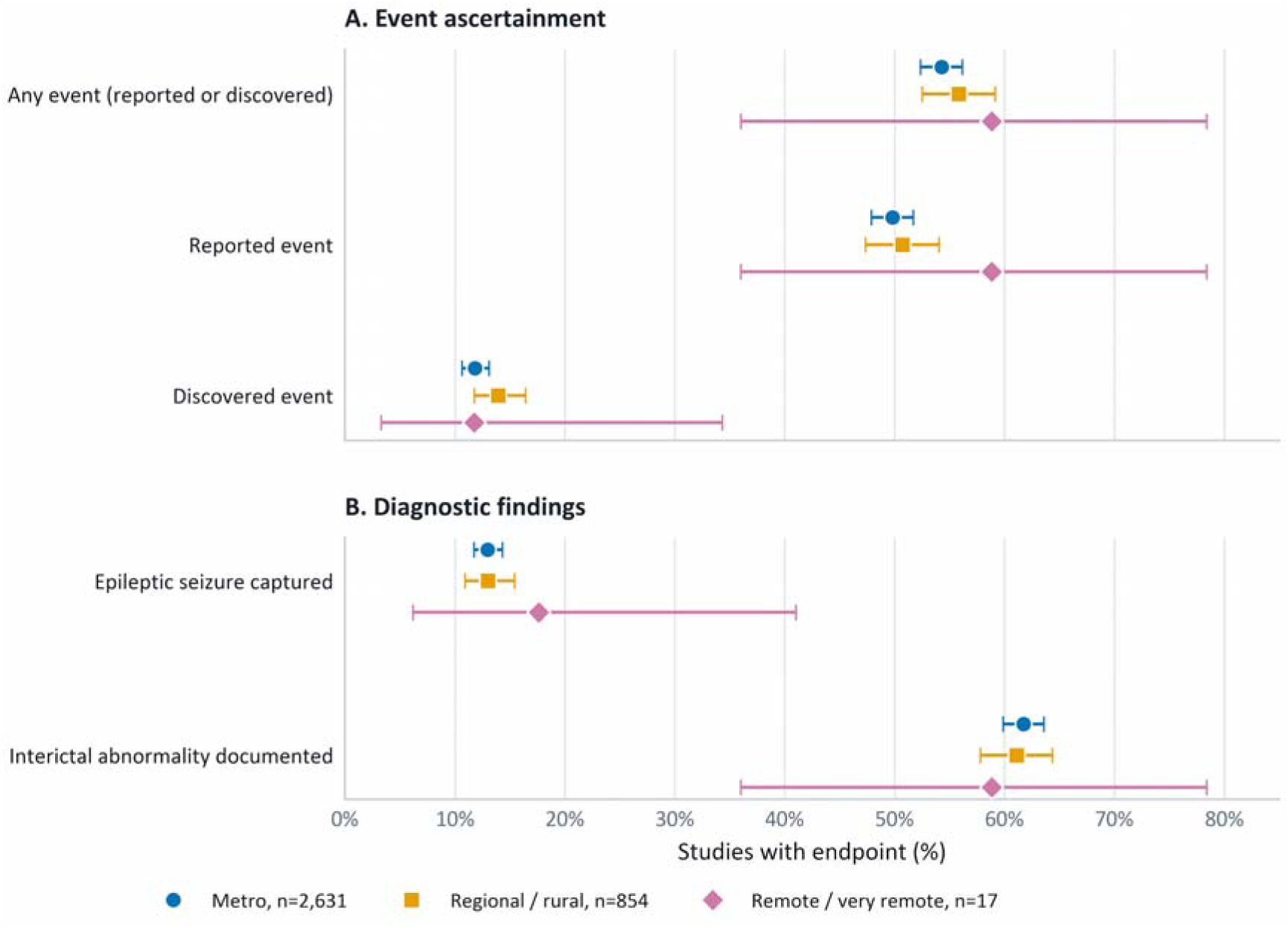
Event ascertainment and diagnostic findings by MMM group. Reported events were logged by patients or carers. Discovered events were identified during clinical review. Seizure and interictal endpoints were derived from neurologist reports. Points show study proportions and whiskers show Wilson 95% confidence intervals. EEG, electroencephalography. MMM, Modified Monash Model.

### Electrode impedance

The median proportion of recorded electrodes below 10 kΩ was 85.7% (IQR 71.4%-90.5%) in metropolitan studies, 85.7% (IQR 71.4%-90.5%) in regional/rural studies, and 81.0% (IQR 66.7%-90.5%) in remote/very remote studies (Figure 3).

**Figure 3.**
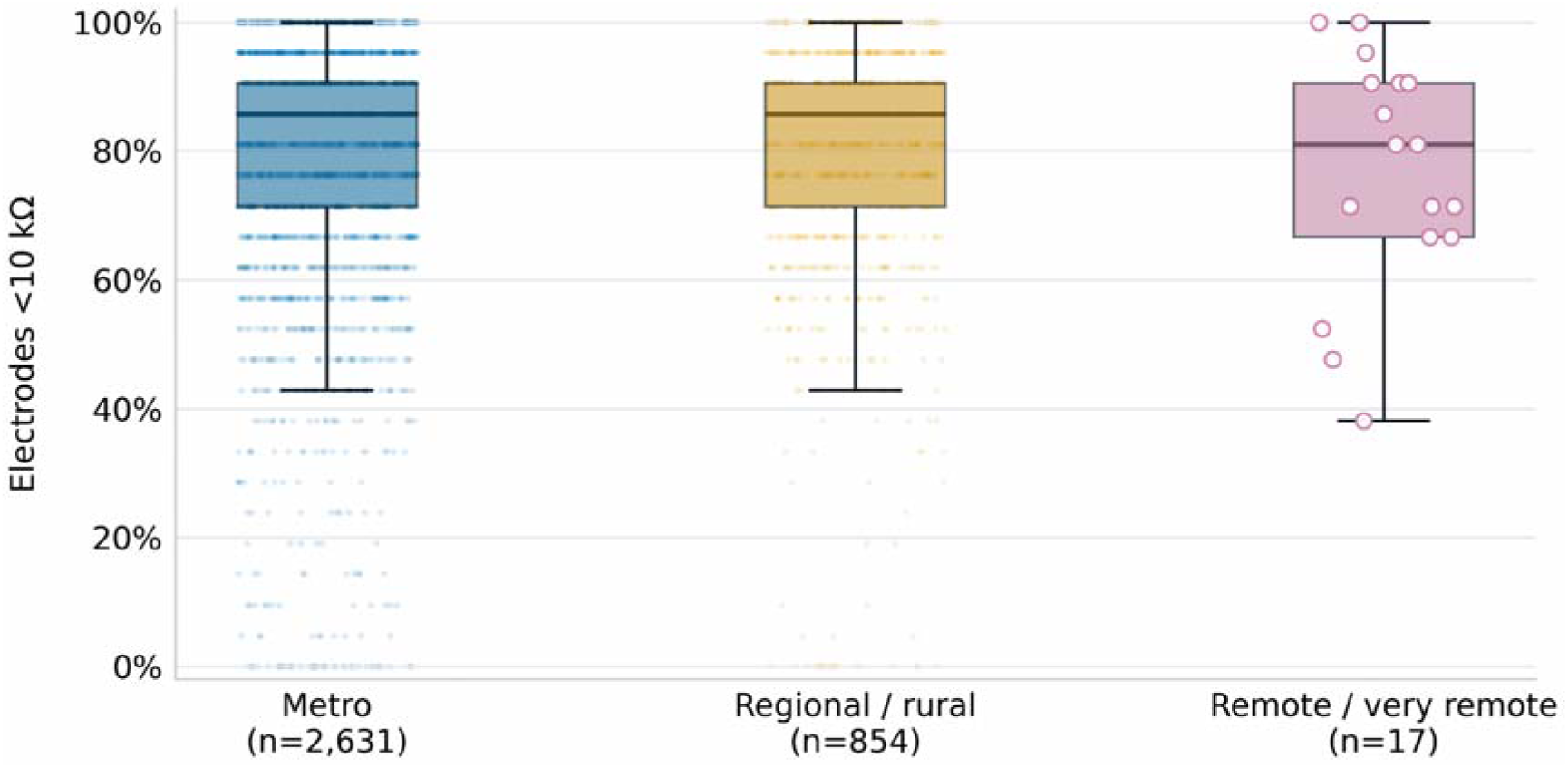
Final-check electrode impedance by MMM group. The outcome is the percentage of recording electrodes below 10 kΩ at the final impedance check. Boxes show the median and interquartile range. Whiskers extend to 1.5 times the interquartile range. Filled points show Metro and regional/rural studies. Open circles show all remote/very remote studies. MMM, Modified Monash Model.

After adjustment for age, sex, recorded epilepsy diagnosis, disability, and recording duration, the estimated mean difference relative to metropolitan studies was 1.48 percentage points for regional/rural studies (95% CI −0.09 to 3.04; P = 0.06) and −1.94 percentage points for remote/very remote studies (95% CI −9.18 to 5.30; P = 0.60). The global MMM association was P = 0.15 (Table S3).

### Recording quality and event observability

A report-documented data-acquisition or event-interpretability issue was present in 107 of 3,502 studies (3.1%). The proportion was 2.9% in metropolitan studies and 3.5% in regional/rural studies. No remote/very remote study had a documented issue. Minor artefact or noise without a major issue was documented in 14.6%, 13.8%, and 17.6% of studies, respectively (Figure 4).

**Figure 4.**
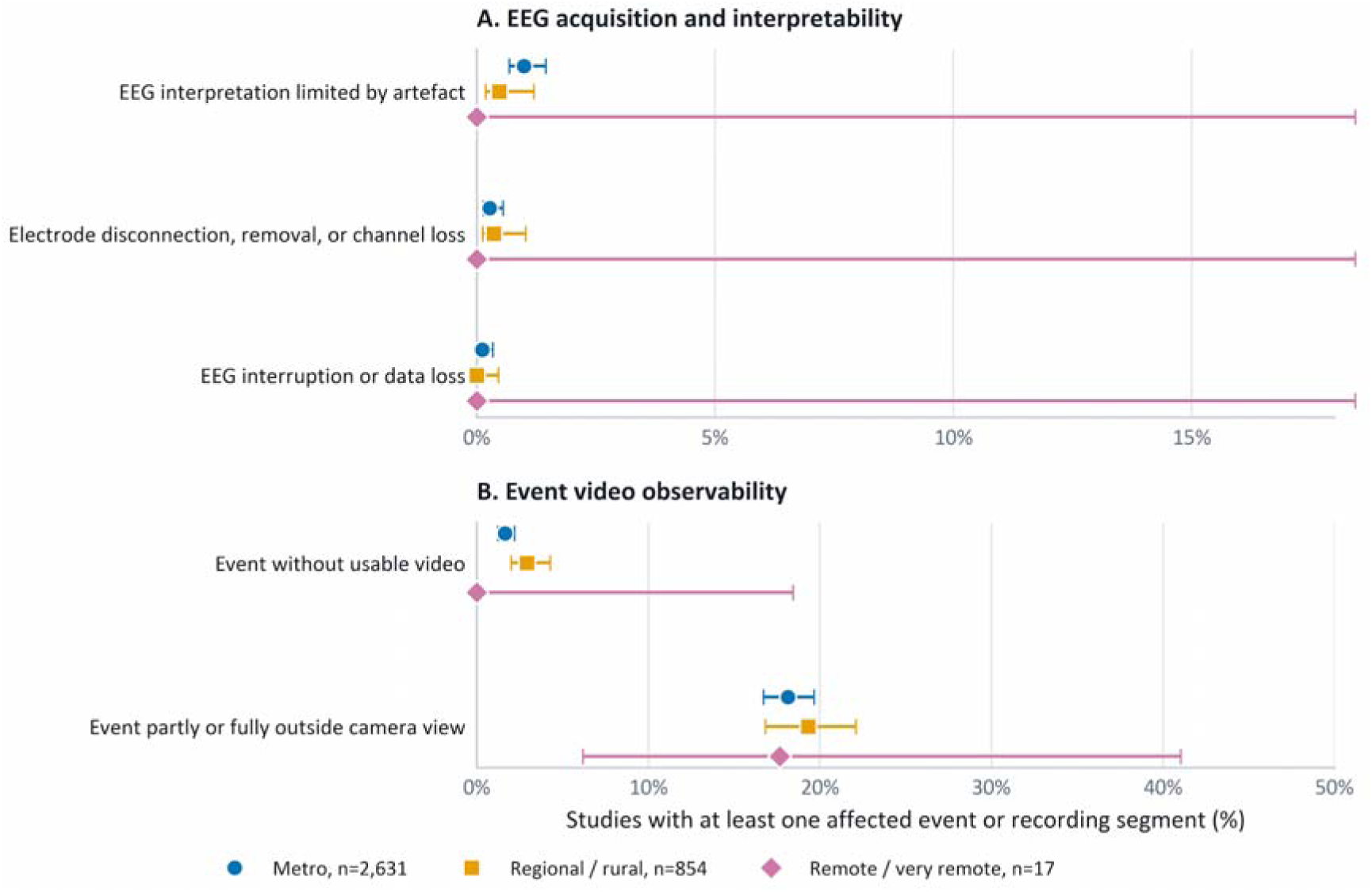
Report-documented recording and video issues by MMM group. A study was positive when at least one event or recording segment met the relevant criterion. EEG artefact includes event-specific obscuration and broader segment-level issues. Event without usable video includes technical, contextual, or intentional unavailability. Points show study proportions and whiskers show Wilson 95% confidence intervals. Panels use different x-axis ranges. EEG, electroencephalography. MMM, Modified Monash Model.

EEG interpretation was documented as limited by artefact in 1.0% of metropolitan studies and 0.5% of regional/rural studies, while electrode disconnection, removal, or channel loss was documented in 0.3% and 0.4%, respectively. Neither issue occurred in remote/very remote studies. EEG interruption or data loss was uncommon in all groups. Exact comparisons provided no evidence of locality differences for these sparse endpoints.

An event without usable video was documented in 43 of 2,631 metropolitan studies (1.6%) and 25 of 854 regional/rural studies (2.9%). No remote/very remote study met this criterion. The unadjusted exact comparison was P = 0.07 (false-discovery-rate q = 0.35). Because the locality data were sparse, a stable two-way-clustered adjusted MMM model was not fitted for this endpoint.

At least one event was partly or fully outside the camera view in 18.1% of metropolitan studies, 19.3% of regional/rural studies, and 17.6% of remote/very remote studies. There was no evidence of an adjusted association with MMM group. The adjusted PR was 1.04 (95% CI 0.89-1.21) for regional/rural studies and 0.97 (95% CI 0.34-2.73) for remote/very remote studies relative to metropolitan studies (global P = 0.87). No locality comparison remained significant after false-discovery-rate correction.

### Geographic distance and area-level disadvantage

Geographic exposure measures were available for all 3,502 studies. We therefore examined assigned-clinic distance and area-level IRSD as continuous exposures in addition to categorical MMM analyses (Figure 5; Table S4).

**Figure 5.**
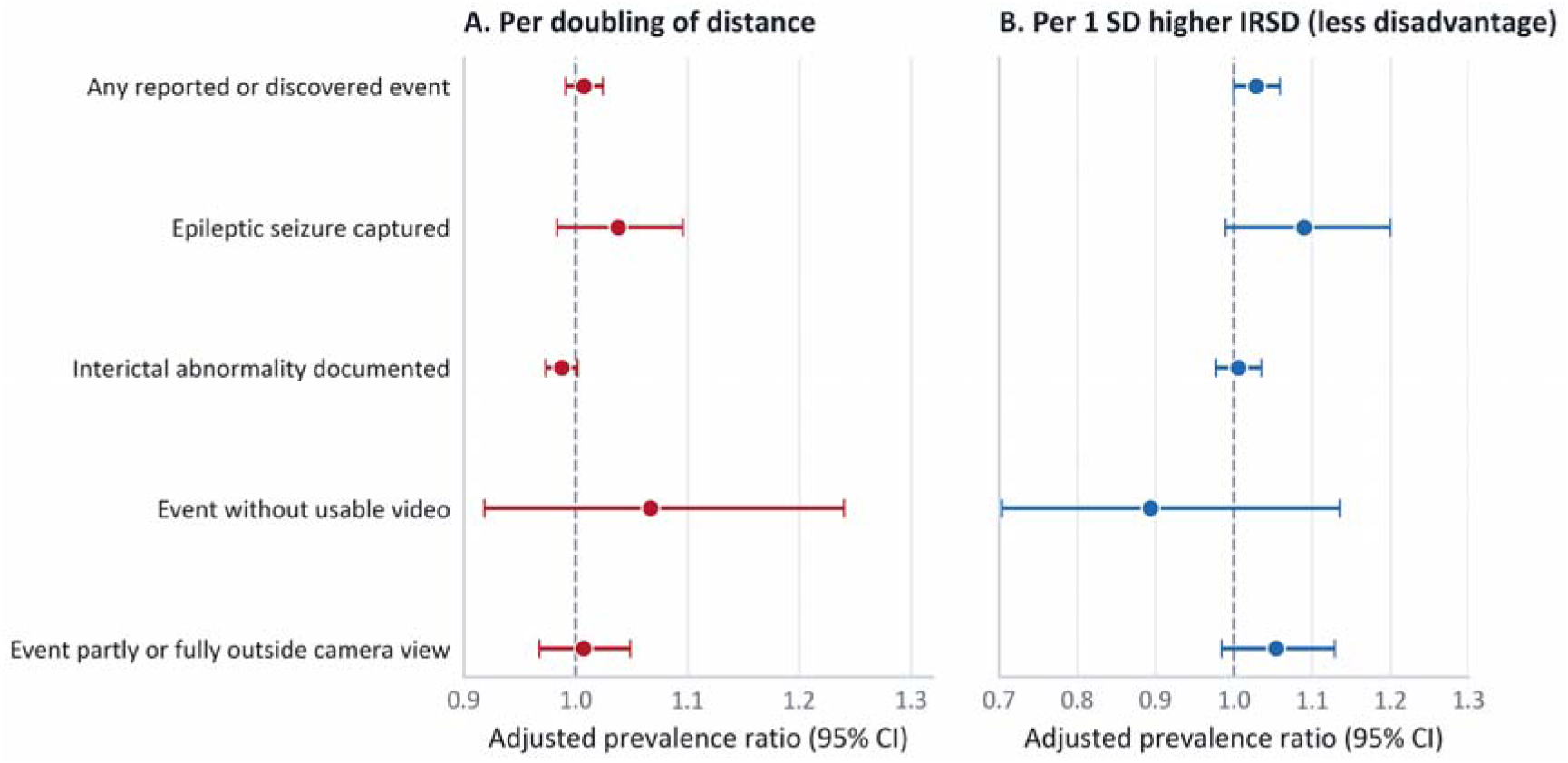
Adjusted associations of geographic exposure with study outcomes. Effects are shown per doubling of assigned-clinic distance and per 1-SD higher IRSD score, representing less disadvantage. Points show adjusted prevalence ratios and whiskers show 95% confidence intervals. Models included both geographic exposures and adjusted for age, sex, recorded epilepsy diagnosis, disability, antiseizure medication count, referral purpose, and recording duration. Covariance was clustered by patient and postcode. Distance is straight-line postcode-centroid distance. Analyses are conditional on receipt of home video-EEG and do not estimate population-level use or unmet access. EEG, electroencephalography; IRSD, Index of Relative Socio-economic Disadvantage; SD, standard deviation.

In models containing assigned-clinic distance and IRSD together, neither exposure showed evidence of an association with any primary outcome after false-discovery-rate correction (Figure 5). For any event, the adjusted prevalence ratio (PR) per doubling of clinic distance was 1.01 (95% CI 0.99-1.02; q = 0.50). The PR per 1-SD higher IRSD score was 1.03 (95% CI 1.00-1.06; q = 0.29). No association remained significant after correction for epileptic seizure capture, interictal abnormality, event without usable video, or event outside the camera view. Final-check impedance was unrelated to distance or IRSD. Results were materially unchanged in clinic-adjusted sensitivity analyses, with no association between MMM group or clinic distance and lower diagnostic yield or poorer technical performance after false-discovery-rate correction (Table S5).

## Discussion

In this Australia-wide cohort of 3,502 home video-EEG studies, home video-EEG was used across 1,112 postcodes, with 24.4% of studies from regional/rural residents. This was broadly similar in magnitude to the proportion of Australians living in Inner/Outer Regional areas, although the population comparator uses the separate Remoteness Area classification and is not directly equivalent to MMM.^18^ Remote/very remote representation was sparse. The data therefore support reduced geographic burden for regional/rural recipients, but do not demonstrate comparable access gains for genuinely remote populations. Assigned clinics were substantially closer than public comprehensive epilepsy centres for many regional/rural recipients, yet there was no evidence that diagnostic yield or technical performance worsened across MMM category, clinic distance, or area-level socioeconomic disadvantage. These findings support distributed home video-EEG as a means of reducing travel burden among service recipients without evidence of lower diagnostic yield or poorer technical performance.

For regional/rural recipients, the assigned clinic was a median 59.3 km closer one way (IQR: 1.2 km farther to 116.6 km closer) than the nearest public comprehensive epilepsy centre and was closer in 68.0% of studies. Because connection and disconnection were clinic-based, this median distance difference corresponds to 237.2 km across four one-way journey legs. Despite this relative advantage, substantial travel remained: median assigned-clinic distance was 73.9 km in regional/rural studies, more than four times the 16.4-km metropolitan median, and 509.6 km in remote/very remote studies. The remote/very remote estimate is based on only 17 studies. These residual distances support further decentralisation through additional regional clinics or self-connection in a setting where regional patients may otherwise travel substantial distances for EEG.^7–9^ Distances are straight-line postcode-centroid estimates rather than observed driving distances, so actual travel is likely to be longer. Referral-to-recording time was also longer outside metropolitan areas: 33.9 versus 25.5 days, an 8.4-day median difference. In an exploratory analysis accounting for assigned clinic, this difference was no longer evident, suggesting that the overall difference largely reflected between-clinic service-delivery differences rather than a consistent within-clinic delay for regional/rural patients. This is consistent with the periodic operation of regional/rural clinic locations. Published waiting times for prolonged video-EEG are heterogeneous. A US claims study reported mean referral-to-video-EEG intervals of 30.6 days for outpatient and 42.5 days for inpatient monitoring, while an epilepsy monitoring unit cohort reported a median 29 days to first admission offer and 83 days to admission.^19,20^ These estimates are not directly comparable because indications, prioritisation, service models, and waiting-time definitions differ. Our interval spans referral to study commencement and should not be interpreted as a pure scheduling queue. More broadly, early attendance at specialist first-seizure clinics has been associated with reduced subsequent all-cause emergency presentations and hospitalisations, underscoring the potential importance of delays across the diagnostic pathway.^21^

A central concern with decentralisation is whether distance from clinic support degrades recording performance. In this cohort, locality and clinic distance were not associated with poorer event ascertainment, seizure or interictal findings, final-check impedance, or video observability, and report-documented major acquisition or interpretability issues were uncommon. This is consistent with earlier findings of stable impedance across remoteness and high EEG/video availability during home recordings.^4,11^ Area-level disadvantage was also unrelated to primary outcomes after multiplicity correction. These null associations do not establish equivalence, but their consistency across independent clinical and technical endpoints is reassuring.

Self-applied systems could reduce the remaining need for clinic attendance. Early adult studies show that patients can perform home EEG over both short and extended periods: EEG@HOME reported estimated six-month continuation of 83%, with good or moderate EEG quality in 8 of 12 participants on subsequent analysis, while HOMEONE achieved successful home recording in 89 of 97 patients and diagnostically adequate recordings in 89.9%.^22–24^ Caregiver-collected multi-night dry-electrode EEG has also been demonstrated in children with Lennox-Gastaut syndrome.^25^ These findings support feasibility in selected patients, while training, adherence, maintenance, and technical support remain important.

For people in regional/rural and remote/very remote areas, self-connection could remove some or all of the four journey legs required by the current service, reducing travel and time away from work or school. Reduced travel may also have environmental co-benefits, although transport mode and emissions were not measured here. The International League Against Epilepsy (ILAE) Climate Change Commission has called for more sustainable epilepsy practice and identified the climate impact of telemedicine versus in-person care as a research priority.^26^ Future evaluations should assess travel burden, cost, and carbon emissions alongside recording performance and implementation outcomes. While self-application will not suit every patient, it has the potential to move the current service interface from a distributed clinic network into the home for a substantial proportion of patients.

Several limitations affect interpretation. This retrospective study evaluated a single service provider with remote technical review, so findings may not generalise to other home EEG or self-application models. Because the cohort included only people who received testing, it cannot estimate population-level use, unmet need, or barriers to access; likewise, the absence of an IRSD association among recipients does not demonstrate equitable access. Only 17 studies were remote/very remote, limiting the ability to draw firm conclusions about this area.

Geographic exposures were based on postcode centroids and straight-line distance, not observed road travel, time, or cost. Public comprehensive epilepsy centres were contextual comparators rather than observed referral alternatives, and the inventory was not age-specific and excluded private providers. Recording-quality outcomes came from neurologist reports rather than systematic re-review, so transient technical issues may be undercaptured. Finally, adjusted null associations cannot establish equivalence. These limitations are partly offset by the cohort’s Australia-wide geographic coverage, the paired geographic comparison, and the consistency of findings across categorical, continuous, and postcode-origin sensitivity analyses.

## Conclusions

Distributed home video-EEG extended prolonged monitoring across a broad Australian geography and substantially reduced the distance to the point of connection for many regional/rural recipients. There was no evidence of lower diagnostic yield or poorer technical performance with remoteness, clinic distance, or area-level disadvantage among those who accessed the service, although regional/rural referral-to-recording intervals were longer overall. Only 17 studies were remote/very remote, so comparable access gains in genuinely remote populations remain uncertain. Future studies should focus on whether decentralisation increases use among underserved populations and reduces the real travel, time, and financial costs of prolonged monitoring. Self-applied systems may further shift the point of access from regional clinics into patients’ homes.

## Author Contributions

Conceptualisation: E.S.N., D.R.F., G.D., M.J.C.; Formal analysis: E.S.N., P.K., M.J.C.; Methodology: E.S.N., E.F.; Software: E.S.N.; Writing - original draft: E.S.N., P.K., E.F., M.J.C.; Writing - review & editing: E.S.N., D.R.F., E.F., G.D., P.K., M.J.C.

## Ethical Publication Statement

Ethics approval was provided by the St Vincent’s Hospital Melbourne Human Research Ethics Committee (project 57392).

## Patient Consent Statement

Written informed consent was provided by patients or, where applicable, a guardian.

## Data Availability Statement

The data underlying this study are not publicly available because they contain potentially sensitive clinical and geographic information. Requests for access to de-identified data may be considered by the corresponding author subject to ethical, institutional, and data-governance requirements.

