## Supporting information for "Geographic use and performance of distributed home video-electroencephalography in Australia"

### Table S1. Relative geographic distance to the assigned EEG clinic and a public epilepsy centre

Panel A. Paired distance summary

| MMM group | Studies, n | Patients, n | Postcodes, n | EEG clinic, median [IQR], km | Public epilepsy centre, median [IQR], km | Paired difference, median [IQR], km | Distance ratio, median [IQR] |
| --- | --- | --- | --- | --- | --- | --- | --- |
| Overall | 3502 | 3457 | 1112 | 19.8 [10.3, 38.3] | 24.5 [9.7, 85.0] | 0.3 [-24.0, 3.9] | 1.0 [0.6, 1.3] |
| Metro | 2631 | 2592 | 739 | 16.4 [9.4, 27.0] | 16.7 [7.9, 33.1] | 1.3 [-1.4, 4.2] | 1.1 [0.9, 1.5] |
| Regional / rural | 854 | 848 | 359 | 73.9 [36.9, 142.0] | 148.8 [91.5, 235.3] | -59.3 [-116.6, 1.2] | 0.6 [0.4, 1.0] |
| Remote / very remote | 17 | 17 | 14 | 509.6 [250.2, 1261.3] | 471.3 [248.5, 1252.2] | -2.2 [-6.6, 1.7] | 1.0 [1.0, 1.0] |

Panel B. Distance-ratio thresholds

| MMM group | EEG clinic ≥2× farther, n (%) | EEG clinic ≥4× farther, n (%) | EEG clinic closer, n (%) |
| --- | --- | --- | --- |
| Overall | 408 (11.7%) | 113 (3.2%) | 1,626 (46.4%) |
| Metro | 402 (15.3%) | 110 (4.2%) | 1,034 (39.3%) |
| Regional / rural | 5 (0.6%) | 2 (0.2%) | 581 (68.0%) |
| Remote / very remote | 1 (5.9%) | 1 (5.9%) | 11 (64.7%) |

Panel C. Exploratory clustered model

| Contrast | Ratio estimate [95% CI] | P value | Global MMM P value |
| --- | --- | --- | --- |
| Regional / rural vs Metro | 0.43 [0.32, 0.59] | <0.001 | <0.001 |
| Remote / very remote vs Metro | 1.17 [0.92, 1.48] | 0.20 |  |
| Per 1 SD higher IRSD | 0.92 [0.83, 1.02] | 0.10 |  |

Panel D. Postcode-origin uncertainty sensitivity

| MMM group | Maximum simulated origin displacement, km | Studies, n | Iterations, n | Median paired difference, km: median [2.5th, 97.5th] | Assigned clinic closer, %: median [2.5th, 97.5th] |
| --- | --- | --- | --- | --- | --- |
| Overall | Metro 2.5; regional/rural 25.0; remote/very remote 75.0 | 3502 | 5000 | 0.7 [0.6, 0.7] | 43.5 [43.1, 44.1] |
| Metro | 2.5 | 2631 | 5000 | 1.6 [1.5, 1.7] | 36.4 [35.8, 37.0] |
| Regional / rural | 25.0 | 854 | 5000 | -57.8 [-59.8, -55.2] | 65.3 [64.4, 66.3] |
| Remote / very remote | 75.0 | 17 | 5000 | -1.8 [-2.0, -1.4] | 58.8 [52.9, 64.7] |

Notes: Paired difference was calculated as assigned EEG clinic minus public epilepsy-centre distance; negative values favour the assigned clinic. Distance ratio was calculated as assigned/public-centre distance; values <1 favour the assigned clinic. Distances are straight-line postcode-centroid measures. AIHW-listed public comprehensive epilepsy centres are geographic comparators, not observed alternative referral destinations. The public inventory excludes private providers and is not age-specific. The exploratory model of log2 assigned/public-centre distance ratio was adjusted for MMM group and IRSD with covariance clustered by patient and postcode. The postcode-origin sensitivity used 5,000 simulations and displaced the assumed patient origin within prespecified maximum radii of 2.5 km for metropolitan, 25 km for regional/rural, and 75 km for remote/very remote studies. AIHW, Australian Institute of Health and Welfare; CI, confidence interval; EEG, electroencephalography; IQR, interquartile range; IRSD, Index of Relative Socio-economic Disadvantage; MMM, Modified Monash Model; SD, standard deviation.

### Table S2. Adjusted associations of MMM group with event, diagnostic, and video endpoints

| Outcome | Contrast | Adjusted PR [95% CI] | P value | Global MMM P value | Global MMM BF01 |
| --- | --- | --- | --- | --- | --- |
| Any reported or discovered event | Regional / rural vs Metro | 1.01 [0.94, 1.08] | 0.73 | 0.85 | 3.05 × 10³ |
|  | Remote / very remote vs Metro | 1.09 [0.77, 1.54] | 0.64 |  |  |
| Any reported event | Regional / rural vs Metro | 1.00 [0.92, 1.08] | 0.96 | 0.60 | 2.54 × 10³ |
|  | Remote / very remote vs Metro | 1.19 [0.85, 1.68] | 0.31 |  |  |
| Any discovered event | Regional / rural vs Metro | 1.16 [0.96, 1.40] | 0.12 | 0.30 | 1.17 × 10³ |
|  | Remote / very remote vs Metro | 0.95 [0.28, 3.20] | 0.94 |  |  |
| Epileptic seizure captured | Regional / rural vs Metro | 0.99 [0.80, 1.21] | 0.90 | 0.80 | 3.07 × 10³ |
|  | Remote / very remote vs Metro | 1.30 [0.59, 2.88] | 0.51 |  |  |
| Interictal abnormality documented | Regional / rural vs Metro | 0.99 [0.93, 1.05] | 0.69 | 0.89 | 3.21 × 10³ |
|  | Remote / very remote vs Metro | 0.95 [0.70, 1.29] | 0.76 |  |  |
| Event without usable video | Sparse locality cells | Not estimated |  |  |  |
| Event partly or fully outside camera view | Regional / rural vs Metro | 1.04 [0.89, 1.21] | 0.60 | 0.87 | 3.08 × 10³ |
|  | Remote / very remote vs Metro | 0.97 [0.34, 2.73] | 0.95 |  |  |

Notes: Metro is the reference group. Modified Poisson models were adjusted for age, sex, recorded epilepsy diagnosis, disability, antiseizure medication count, referral purpose, and recording duration, with covariance clustered by patient and postcode. The event-without-usable-video endpoint had sparse locality cells and was not estimated in the adjusted MMM model. Global MMM P values and BF01 are shown once per outcome. BF01 uses an unclustered covariate-matched logistic-model Bayesian information criterion (BIC) approximation. PR, prevalence ratio; CI, confidence interval; MMM, Modified Monash Model; BF01, Bayes factor in favour of the model without an MMM association.

### Table S3. Adjusted associations with final-check electrode impedance

| Exposure | Contrast or unit | Adjusted difference [95% CI], percentage points | P value | Global MMM P value | Global MMM BF01 | Studies / patients, n |
| --- | --- | --- | --- | --- | --- | --- |
| MMM group | Regional / rural vs Metro | 1.48 [-0.09, 3.04] | 0.06 | 0.15 | 471.65 | 3502 / 3457 |
| MMM group | Remote / very remote vs Metro | -1.94 [-9.18, 5.30] | 0.60 |  |  |  |
| Distance to assigned EEG clinic | Per doubling of distance | 0.12 [-0.22, 0.46] | 0.50 |  |  | 3502 / 3457 |
| Area-level IRSD | Per 1 SD higher score (less disadvantage) | 0.25 [-0.45, 0.96] | 0.48 |  |  | 3502 / 3457 |

Notes: The MMM model was adjusted for age, sex, recorded epilepsy diagnosis, disability, and recording duration. Distance and IRSD models contained both geographic exposures and the same clinical covariates. Covariance was clustered by patient and postcode. Estimates are percentage-point differences. Distance is interpreted per doubling; IRSD is interpreted per 1-SD higher score, representing less area-level disadvantage. BF01 uses an unclustered covariate-matched linear-model BIC approximation. BF01, Bayes factor in favour of the model without an MMM association; BIC, Bayesian information criterion; CI, confidence interval; EEG, electroencephalography; IRSD, Index of Relative Socio-economic Disadvantage; MMM, Modified Monash Model; SD, standard deviation.

### Table S4. Geographic exposure, referral-to-recording interval, and adjusted associations with geographic distance and area-level disadvantage

Panel A. Geographic exposure and referral-to-recording distributions

| MMM group | Studies, n | Distance to assigned EEG clinic, median [IQR], km | IRSD decile, median [IQR] |
| --- | --- | --- | --- |
| Overall | 3502 | 19.8 [10.3, 38.3] | 6.0 [3.0, 8.0] |
| Metro | 2631 | 16.4 [9.4, 27.0] | 7.0 [4.0, 9.0] |
| Regional / rural | 854 | 73.9 [36.9, 142.0] | 4.0 [2.0, 6.0] |
| Remote / very remote | 17 | 509.6 [250.2, 1261.3] | 5.0 [3.0, 5.0] |

Referral-to-recording interval was available for all 3,502 studies. Median intervals were 27.3 days (IQR 14.3-48.5) overall, 25.5 days (IQR 13.5-45.5) in metropolitan studies, 33.9 days (IQR 17.8-62.3) in regional/rural studies, and 34.6 days (IQR 24.5-73.7) in remote/very remote studies. The interval differed by MMM group (Kruskal-Wallis P < 0.001; false-discovery-rate q = 0.001; BF01 < 0.01). It was calculated from linked referral date to recording start.

Panel B. Adjusted associations

| Outcome | Exposure | Unit | Adjusted PR [95% CI] | P value |
| --- | --- | --- | --- | --- |
| Any reported or discovered event | Distance to assigned EEG clinic | Per doubling of distance | 1.01 [0.99, 1.02] | 0.38 |
| Any reported or discovered event | Area-level IRSD | Per 1 SD higher score (less disadvantage) | 1.03 [1.00, 1.06] | 0.05 |
| Epileptic seizure captured | Distance to assigned EEG clinic | Per doubling of distance | 1.04 [0.98, 1.10] | 0.18 |
| Epileptic seizure captured | Area-level IRSD | Per 1 SD higher score (less disadvantage) | 1.09 [0.99, 1.20] | 0.08 |
| Interictal abnormality documented | Distance to assigned EEG clinic | Per doubling of distance | 0.99 [0.97, 1.00] | 0.09 |
| Interictal abnormality documented | Area-level IRSD | Per 1 SD higher score (less disadvantage) | 1.01 [0.98, 1.04] | 0.69 |
| Event without usable video | Distance to assigned EEG clinic | Per doubling of distance | 1.07 [0.92, 1.24] | 0.40 |
| Event without usable video | Area-level IRSD | Per 1 SD higher score (less disadvantage) | 0.89 [0.70, 1.14] | 0.36 |
| Event partly or fully outside camera view | Distance to assigned EEG clinic | Per doubling of distance | 1.01 [0.97, 1.05] | 0.73 |
| Event partly or fully outside camera view | Area-level IRSD | Per 1 SD higher score (less disadvantage) | 1.05 [0.98, 1.13] | 0.13 |

Notes: Distance and IRSD were entered together. Modified Poisson models were adjusted for age, sex, recorded epilepsy diagnosis, disability, antiseizure medication count, referral purpose, and recording duration, with covariance clustered by patient and postcode. Distance is interpreted per doubling; IRSD is interpreted per 1-SD higher score, representing less area-level disadvantage. CI, confidence interval; EEG, electroencephalography; IQR, interquartile range; IRSD, Index of Relative Socio-economic Disadvantage; MMM, Modified Monash Model; PR, prevalence ratio; SD, standard deviation.

### Table S5. Sensitivity analyses accounting for assigned clinic

Panel A. Clinic-locality overlap

| Clinic-overlap measure | Clinics, n | Regional / rural studies, n | Remote / very remote studies, n |
| --- | --- | --- | --- |
| All assigned clinics | 24 | 854 | 17 |
| Clinics with metropolitan and non-metropolitan observations | 17 | 597 | 14 |

Panel B. Clinic-adjusted MMM associations

| Outcome | Contrast | Clinic-adjusted estimate [95% CI] | P value | FDR q |
| --- | --- | --- | --- | --- |
| Any reported or discovered event | Regional / rural vs Metro | 1.05 [0.96, 1.14] | 0.28 | 0.92 |
| Any reported or discovered event | Remote / very remote vs Metro | 1.08 [0.75, 1.56] | 0.67 | 0.92 |
| Epileptic seizure captured | Regional / rural vs Metro | 1.09 [0.85, 1.41] | 0.49 | 0.92 |
| Epileptic seizure captured | Remote / very remote vs Metro | 1.28 [0.57, 2.87] | 0.55 | 0.92 |
| Interictal abnormality documented | Regional / rural vs Metro | 1.00 [0.93, 1.08] | 0.92 | 0.92 |
| Interictal abnormality documented | Remote / very remote vs Metro | 0.95 [0.70, 1.28] | 0.72 | 0.92 |
| Event without usable video | Sparse locality cells | Not estimated |  |  |
| Event partly or fully outside camera view | Regional / rural vs Metro | 1.20 [1.00, 1.44] | 0.05 | 0.41 |
| Event partly or fully outside camera view | Remote / very remote vs Metro | 0.90 [0.32, 2.51] | 0.84 | 0.92 |
| Final-check impedance, percentage-point difference | Regional / rural vs Metro | 0.34 [-1.67, 2.34] | 0.74 | 0.74 |
| Final-check impedance, percentage-point difference | Remote / very remote vs Metro | -1.96 [-9.46, 5.53] | 0.61 | 0.74 |

Panel C. Clinic-adjusted continuous geographic associations

| Outcome | Exposure | Unit | Clinic-adjusted estimate [95% CI] | P value / FDR q |
| --- | --- | --- | --- | --- |
| Any reported or discovered event | Distance to assigned EEG clinic | Per doubling of distance | 1.01 [0.99, 1.02] | 0.53 / 0.66 |
| Any reported or discovered event | Area-level IRSD | Per 1 SD higher score | 1.03 [1.00, 1.06] | 0.09 / 0.29 |
| Epileptic seizure captured | Distance to assigned EEG clinic | Per doubling of distance | 1.04 [0.98, 1.09] | 0.23 / 0.38 |
| Epileptic seizure captured | Area-level IRSD | Per 1 SD higher score | 1.09 [0.98, 1.20] | 0.11 / 0.29 |
| Interictal abnormality documented | Distance to assigned EEG clinic | Per doubling of distance | 0.99 [0.97, 1.00] | 0.06 / 0.29 |
| Interictal abnormality documented | Area-level IRSD | Per 1 SD higher score | 1.00 [0.97, 1.03] | 1.00 / 1.00 |
| Event without usable video | Distance to assigned EEG clinic | Per doubling of distance | 1.06 [0.91, 1.22] | 0.46 / 0.66 |
| Event without usable video | Area-level IRSD | Per 1 SD higher score | 0.81 [0.64, 1.03] | 0.08 / 0.29 |
| Event partly or fully outside camera view | Distance to assigned EEG clinic | Per doubling of distance | 1.01 [0.97, 1.05] | 0.73 / 0.81 |
| Event partly or fully outside camera view | Area-level IRSD | Per 1 SD higher score | 1.05 [0.98, 1.13] | 0.17 / 0.34 |
| Final-check impedance, percentage-point difference | Distance to assigned EEG clinic | Per doubling of distance | 0.11 [-0.25, 0.47] | 0.55 / 0.55 |
| Final-check impedance, percentage-point difference | Area-level IRSD | Per 1 SD higher score | 0.44 [-0.28, 1.17] | 0.23 / 0.46 |

Panel D. Exploratory referral-to-recording analysis

| Analysis / stratum | Studies, n | Median [IQR], days or ratio [95% CI] | P value |
| --- | --- | --- | --- |
| Metropolitan residents, metropolitan clinic | 2,627 | 25.5 [13.5, 45.5] days |  |
| Regional / rural residents, metropolitan clinic | 500 | 26.5 [14.2, 46.9] days |  |
| Regional / rural residents, regional / rural clinic | 354 | 49.4 [26.5, 85.1] days |  |
| Clinic-adjusted regional / rural vs Metro | 3,502 | 0.99 [0.91, 1.07] | 0.81 |
| Clinic-adjusted remote / very remote vs Metro | 3,502 | 1.15 [0.72, 1.83] | 0.55 |

Notes: Sensitivity analyses repeated the primary adjusted models with assigned clinic included as a categorical fixed effect; patient- and postcode-clustered covariance was otherwise retained. Clinic-locality overlap was summarised because clinic and residential locality were related. Modified Poisson models retained the clinical covariates used in the primary analyses. Final-check impedance estimates are percentage-point differences; other binary-outcome estimates are prevalence ratios. False-discovery-rate q values were calculated within the corresponding clinic-adjusted sensitivity families. The event-without-usable-video endpoint had sparse locality cells and was not estimated in the clinic-adjusted categorical MMM model. Referral-to-recording interval was examined in an exploratory model of log(days + 1). Regional/rural clinic locations operated periodically rather than as continuously staffed permanent sites. CI, confidence interval; EEG, electroencephalography; IRSD, Index of Relative Socio-economic Disadvantage; MMM, Modified Monash Model; PR, prevalence ratio; SD, standard deviation.
